# Large-scale audiometric phenotyping identifies distinct genes and pathways involved in hearing loss subtypes

**DOI:** 10.1101/2025.01.14.24318673

**Authors:** Samah Ahmed, Kenneth I. Vaden, Darren Leitao, Judy R. Dubno, Britt I. Drögemöller

## Abstract

Age-related hearing loss affects one-third of the population over 65 years. However, the diverse pathologies underlying these heterogenous phenotypes complicate genetic studies. To overcome challenges associated with accurate phenotyping for older adults with hearing loss, we applied computational phenotyping approaches based on audiometrically measured hearing loss. This novel phenotyping strategy uncovered distinct genetic variants associated with sensory and metabolic hearing loss. Sex-stratified analyses of these sexually dimorphic hearing loss phenotypes revealed a novel locus of relevance to sensory hearing loss in males, but not females. Enrichment analyses revealed that genes involved in frontotemporal dementia were implicated in metabolic hearing loss, while genes relating to sensory processing of sound by hair cells were implicated in sensory hearing loss. Our study has enhanced our understanding of these two distinct hearing loss phenotypes, representing the first step in the development of more precise treatments for these pathologically distinct hearing loss phenotypes.

## 1. Introduction

Hearing loss affects more than 1.5 billion people around the world, with the costs attributed to unaddressed hearing loss amounting to US$980 billion per year.^1^ In older adults, hearing loss is the most common sensory impairment. In fact, almost one-third of the population over 65 experience difficulties hearing,^2^ with these numbers steadily increasing with an aging population.^3^ The presence of hearing loss is associated with loneliness, stigmatization, depression, and communication difficulties, significantly impacting the quality of life of older individuals.^4^ As age-related hearing loss (ARHL) is a gradually progressive phenotype, early detection before the onset of severe hearing loss is challenging.^5^ For these reasons, improved strategies for the early and precise detection of hearing loss are crucial to guide optimal treatment and management strategies.

ARHL is a multifactorial disorder where genetic predisposition, together with external factors (e.g., noise exposure, aging and certain diseases and ototoxic drugs), contribute to the development of loss of hearing.^6^ In line with this complexity, diverse hearing loss phenotypes involve distinct pathologies.^7^ For example, ARHL that is consistent with excessive exposure to noise, referred to as sensory hearing loss, is typically accompanied by damage to, or death of, cochlear hair cells. These cells are responsible for transforming sound vibrations into electrical signals at specific regions of the cochlea.^8^ In contrast, metabolic hearing loss, is typically associated with deterioration of cells in the stria vascularis, the structure that maintains the endocochlear potential in the inner ear.^9^ These distinct biological underpinnings are further complicated through sex-related differences, with males at higher risk for sensory hearing loss.^10^

While environmental factors play an important role in ARHL, twin and family-based studies have reported that the heritability of ARHL ranges between 30-70%.^11–14^ Genome-wide association studies (GWAS) have uncovered more than 150 loci that are associated with ARHL. Unfortunately, these studies have relied predominantly on self-reported hearing loss.^15–17^ Given the heterogeneity of hearing loss, it is likely that the heritability and genetics underlying the diverse pathologies associated with ARHL will differ. Therefore, understanding the genetic pathways involved in the metabolic and sensory components of ARHL is critical for developing precise and targeted therapies. These differences should also be explored in the context of sex differences, as evidence suggests that sexual dimorphisms related to ARHL may not only reflect differences in environmental risk factors, but also differences in underlying biological processes.^18^

To address these shortcomings, we applied mathematical approaches to estimate metabolic and sensory components of ARHL using audiogram measurements available through the Canadian Longitudinal Study on Aging (CLSA).^19^ Using these novel approaches in combination with comprehensive GWAS approaches, which included the X chromosome and sex-stratified analyses, our study revealed for the first time that different genetic variants, genes and cellular mechanisms underlie metabolic and sensory hearing loss, as well as the observed sex-differences in ARHL.

## 2. Results

### 2.1. Model-based phenotyping to estimate metabolic and sensory hearing loss

Genotype data was available for 26,622 samples included in the CLSA cohort. Following the genotype and phenotype filtering process detailed in **Supplementary Figures 1 and 2**, a total of 20,332 samples were selected for further analyses. In line with previously published work by Vaden *et al*.^10^, examination of the individual metabolic and sensory estimates revealed that metabolic hearing loss estimates increased more substantially with age compared to sensory estimates, while males showed higher sensory hearing loss compared to females (**Figure 1**).

**Figure 1:**
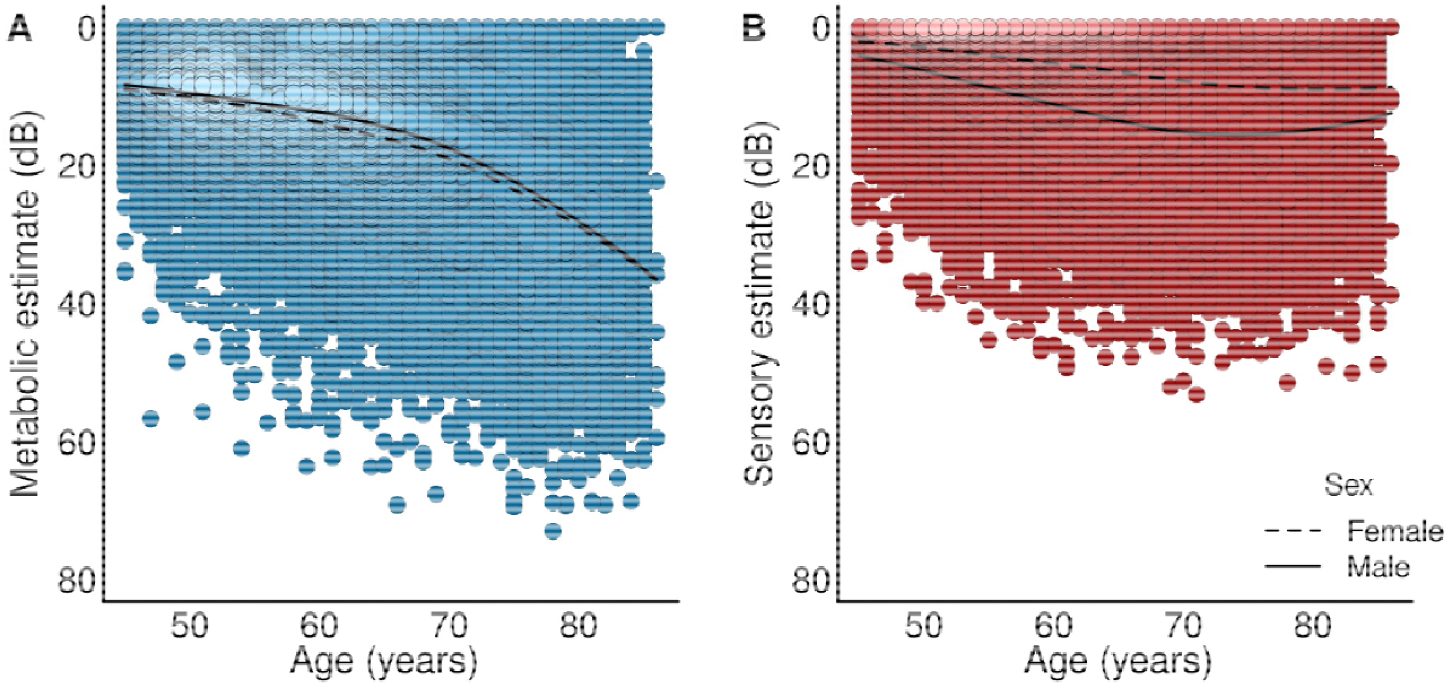
Comparison of metabolic and sensory estimates for the better hearing ear for the same participants. **(A)** Metabolic estimates increase with age, with no substantial difference between males and females. **(B)** Sensory estimates were comparatively less dependent on age and males showed higher sensory hearing loss estimates compared to females. dB: Decibels.

### 2.2. Association of demographic and clinical covariates with metabolic and sensory hearing loss

Analysis of self-reported clinical covariates revealed that older age, sex, diabetes and hypertension were associated with both metabolic and sensory hearing estimates. Osteoporosis and higher sensory hearing estimates were only associated with metabolic hearing estimates. Kidney disease, military service and higher metabolic hearing estimates were only associated with sensory hearing estimates. Variables selected by forward regression analysis are shown in **Table 1**.

**Table 1:**
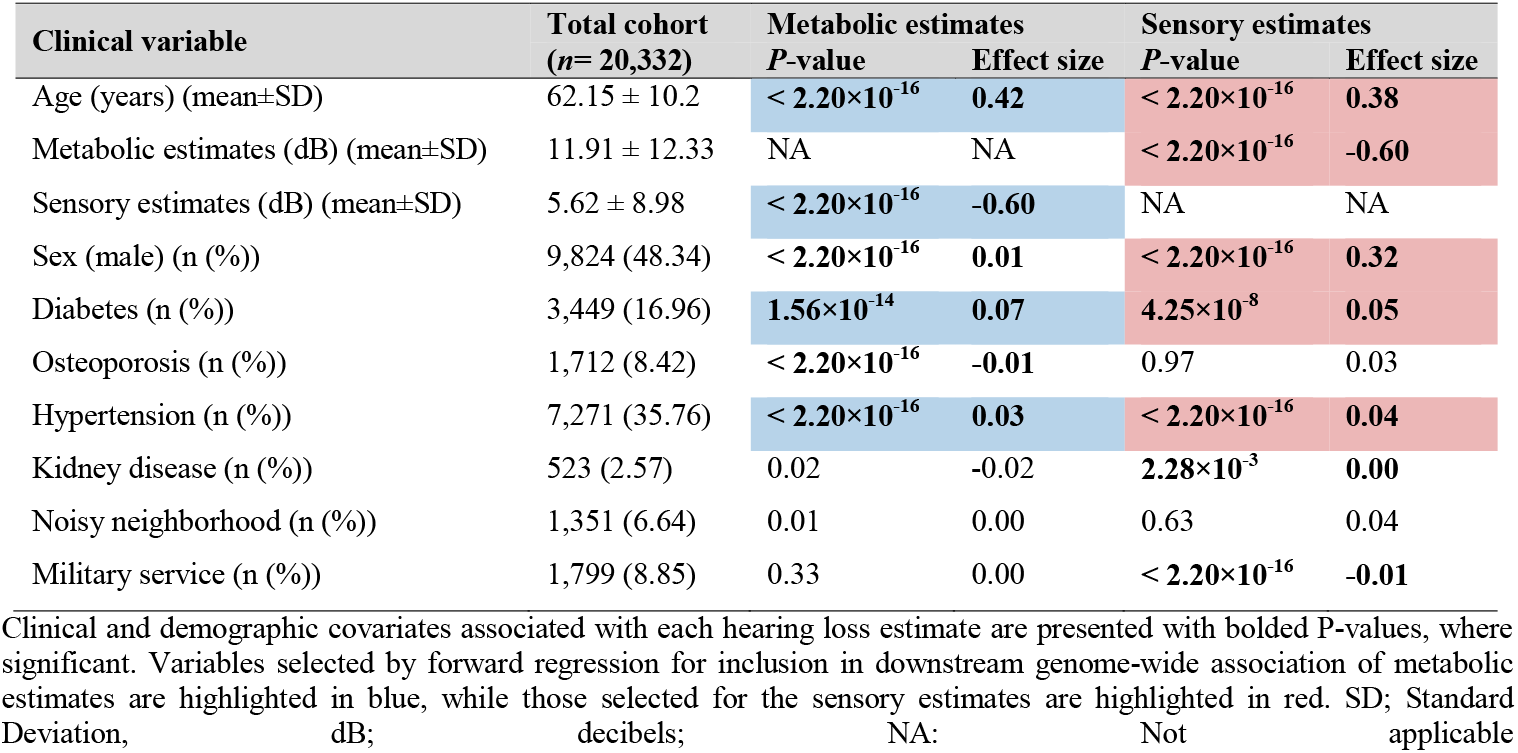
Association between clinical and demographic variables and metabolic and sensory hearing loss.

| Clinical variable | Total cohort<br>(n= 20,332) | Metabolic estimates |  | Sensory estimates |  |
| --- | --- | --- | --- | --- | --- |
|  |  | P-value | Effect size | P-value | Effect size |
| Age (years) (mean±SD) | 62.15 ± 10.2 | **< 2.20×10<sup>-16</sup>** | **0.42** | **< 2.20×10<sup>-16</sup>** | **0.38** |
| Metabolic estimates (dB) (mean±SD) | 11.91 ± 12.33 | NA | NA | **< 2.20×10<sup>-16</sup>** | **-0.60** |
| Sensory estimates (dB) (mean±SD) | 5.62 ± 8.98 | **< 2.20×10<sup>-16</sup>** | **-0.60** | NA | NA |
| Sex (male) (n (%)) | 9,824 (48.34) | **< 2.20×10<sup>-16</sup>** | **0.01** | **< 2.20×10<sup>-16</sup>** | **0.32** |
| Diabetes (n (%)) | 3,449 (16.96) | **1.56×10<sup>-14</sup>** | **0.07** | **4.25×10<sup>-8</sup>** | **0.05** |
| Osteoporosis (n (%)) | 1,712 (8.42) | **< 2.20×10<sup>-16</sup>** | **-0.01** | 0.97 | 0.03 |
| Hypertension (n (%)) | 7,271 (35.76) | **< 2.20×10<sup>-16</sup>** | **0.03** | **< 2.20×10<sup>-16</sup>** | **0.04** |
| Kidney disease (n (%)) | 523 (2.57) | 0.02 | -0.02 | **2.28×10<sup>-3</sup>** | **0.00** |
| Noisy neighborhood (n (%)) | 1,351 (6.64) | 0.01 | 0.00 | 0.63 | 0.04 |
| Military service (n (%)) | 1,799 (8.85) | 0.33 | 0.00 | **< 2.20×10<sup>-16</sup>** | **-0.01** |

Clinical and demographic covariates associated with each hearing loss estimate are presented with bolded P-values, where significant. Variables selected by forward regression for inclusion in downstream genome-wide association of metabolic estimates are highlighted in blue, while those selected for the sensory estimates are highlighted in red. SD; Standard Deviation, dB; decibels; NA: Not applicable

### 2.3. Variants and genes associated with metabolic hearing loss

Genome-wide association analyses identified two genomic risk loci that were associated with metabolic hearing loss, but not sensory hearing loss (*P*<5×10^−8^; **Figure 2A**). Within the first risk locus on chromosome 5, fine-mapping using SuSiE-inf and FINEMAP-inf did not identify any variants in this region with posterior inclusion probability (PIP)>0.5. However, PolyFun assigned a PIP score nearing this predefined threshold (PIP=0.48) for rs6453022, the lead variant in this locus (*P*=2.67×10^−9^; **Supplementary Figure 3**). Annotation analyses revealed that this variant is a missense variant (p.Pro284Gln) in *ARHGEF28*, a member of the Rho guanine nucleotide exchange factor family.^16^ Investigation of this region revealed that rs6453022 is in high linkage disequilibrium (LD) with several other variants (**Supplementary Figure 4A**), likely complicating fine-mapping of this region and leading to the assignment of a relatively low PIP score for the lead variant.

**Figure 2:**
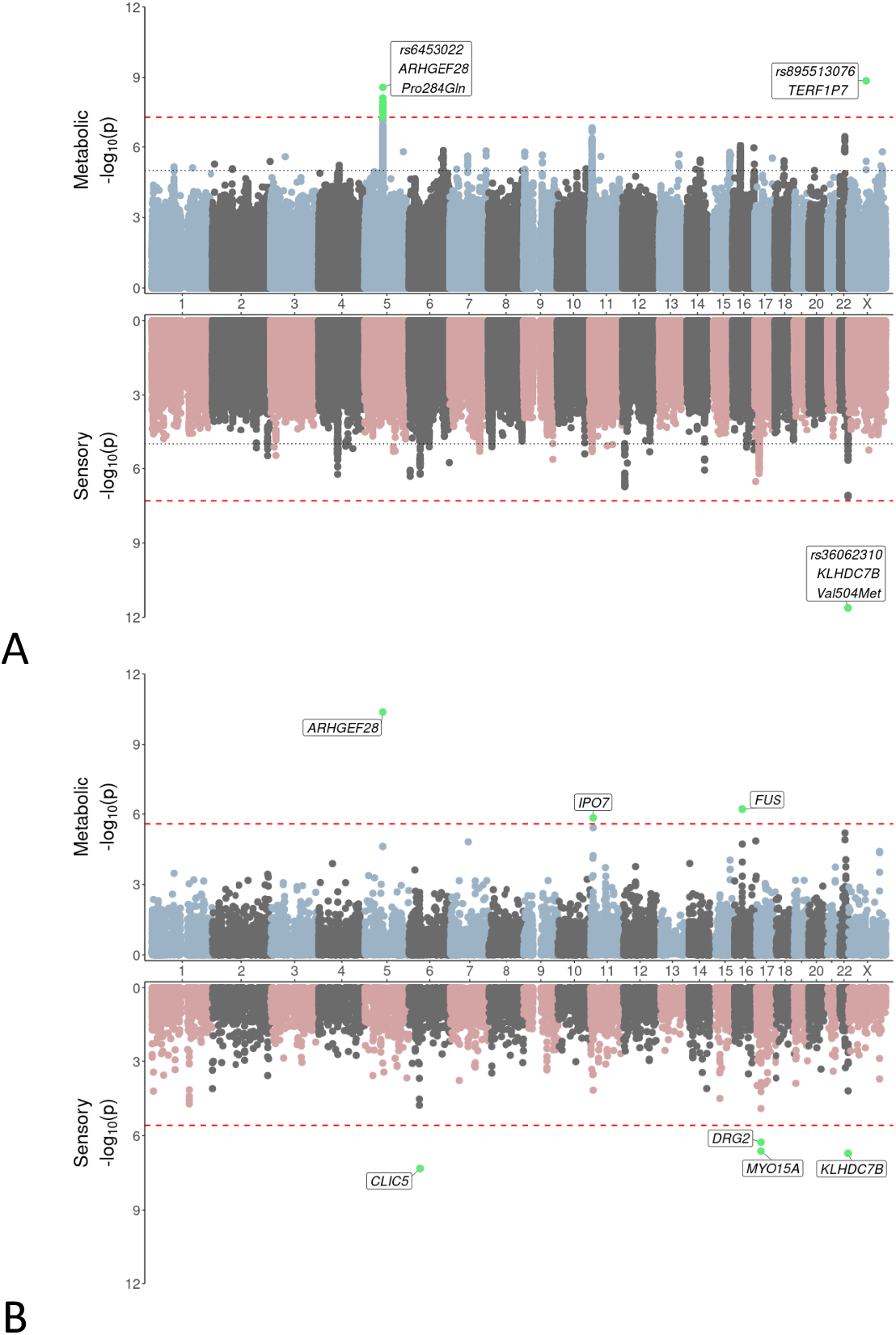
Association between genetic variants and metabolic and sensory hearing loss. The results for metabolic and sensory phenotypes are shown in blue and red in the upper and lower panels, respectively. **(A) Miami plot of genome-wide association study analyses**. The dashed red line represents genome-wide significance (*P*<5×10^−8^). Significant variants are highlighted in green and top variants are labeled. **(B) Miami plot of the gene-based analysis**. The red dashed line represents genome-wide significance (2.57×10^−6^). Genes significantly associated with metabolic or sensory estimates are highlighted in green and labeled.

Closer investigation of this region revealed the presence of a second missense variant in *ARHGEF28*, rs7714670 (p.Trp225Arg) that was also associated with metabolic hearing loss (*P*=2.51×10^−8^), which was in high, but incomplete LD, with the lead variant (D’=1.0, r^2^=0.85). While these coding variants both result in *ARHGEF28* amino acid changes, the impact of the variants on protein function appears to be subtle, with both variants predicted to be benign (SIFT) and tolerated (PolyPhen) for all *ARHGEF28* transcripts, with relatively low CADD scores (rs6453022: CADD=5.42; rs7714670: CADD=9.01). The occurrence of multiple putative causal variants in this region was further supported by our MAGMA gene-based analyses, which revealed an even stronger association between *ARHGEF28* and metabolic hearing loss (MAGMA gene-based *P*=4.17 x10^−11^ vs. GWAS variant-based *P*=2.67×10^−9^). In addition to the association between ARHGEF28 and metabolic hearing loss that was uncovered in the gene-based analyses, a further two autosomal genes were uncovered in these analyses (**Figure 2B**) — FUS, located on chromosome 16 (P=6.17×10-7), coding for an RNA binding protein, and IPO7, located on chromosome 11 (P=1.45×10-6), coding for a nuclear import protein.

The second locus that was significantly associated with metabolic hearing loss was uncovered through our XWAS analyses. The lead variant, rs895513076 (*P*=1.39×10^−9^; model: complete X-inactivation) with PIP>0.9, was not predicted to alter the function of any protein coding genes, but was found to occur 2,423bp upstream of *TERF1P7*, a processed pseudogene. Although gene-based analysis of the X chromosome did not reveal any genes reaching our predefined genome-wide significance threshold, the association between *BCORL1*, a transcriptional corepressor, and metabolic hearing loss trended towards significance (*P*=3.9×10^−5^). No variants reached genome-wide significance in any of our sex-stratified analysis for the metabolic phenotype.

### 2.4. Variants and genes associated with sensory hearing loss

Our GWAS of sensory hearing loss uncovered a genomic risk locus on chromosome 22 that was significantly associated with sensory hearing loss (*P*=2.37×10^−12^; **Figure 2A** and **Supplementary Figure 4B**), but not metabolic hearing loss (*P*=0.001). Fine-mapping of this region revealed that the top variant, rs36062310, had a high PIP score (PIP>0.9) and is also a missense variant (p. Val504Met; ENST00000395676.4/Val1145Met; ENST00000648057.3) in *KLHDC7B*, a gene that has been implicated in toxin-mediated ER stress and apoptosis.^20^ This variant has a relatively high CADD score (22.8) and is predicted to have a deleterious (SIFT) and possibly damaging (PolyPhen) effect on the longer protein coding *KLHDC7B* transcript (ENST00000648057.3).

Gene-based analyses revealed that in addition to the association between *KLHDC7B* and sensory hearing loss (*P*=1.94×10^−7^), a further three autosomal genes were associated with sensory hearing loss **(Figure 2B)**. This includes two known autosomal recessive nonsyndromic hearing loss genes (https://hereditaryhearingloss.org/recessive), *CLIC5* on chromosome 6 (*P*=4.74×10^−8^) and *MYO15A* on chromosome 17 (*P=*2.38×10^−7^), along with *DRG2* (*P=*5.46×10^−7^), a GTP-binding protein, located close to *MYO15A* on chromosome 17. While *CLIC5*, located on chromosome 6, was associated with sensory hearing loss, no variants or genes in the *HLA* region reached genome-wide significance for either metabolic or the sensory phenotypes.

Our sex-stratified analyses revealed that rs36062310 (p.Val1145Met; *KLHDC7B*), the sensory hearing loss-associated variant, was more significantly associated with this phenotype in females compared to males (combined *P*=2.37×10^−12^; females *P*=8.01×10^−9^; males *P*=4.00 ×10^−5^). Additionally, we uncovered a new association between rs72660110 on chromosome 8 and sensory hearing loss. This variant is located proximal to *SULF1*, a gene that has been shown to play an important role in inner ear development.^21^ This variant was significantly associated with the sensory phenotype in males (*P*=2.88×10^−8^), but not females (*P*=0.44). Fine-mapping of this region revealed that although rs72660110 is the most significant variant, it is not likely to be the causal variant (SuSiE-inf PIP=0 and FINEMAP-inf PIP=0.003). While three other variants in this locus all assigned PIP scores of 1 by both SuSiE-inf and FINEMAP-inf, none of these variants were identified as likely causal by PolyFun (PIP<0.5), with rs72660104, an intronic variant, receiving the highest score in this region (PIP=0.289). A list of the potential causal variants is shown in **Table 2**.

**Table 2:**
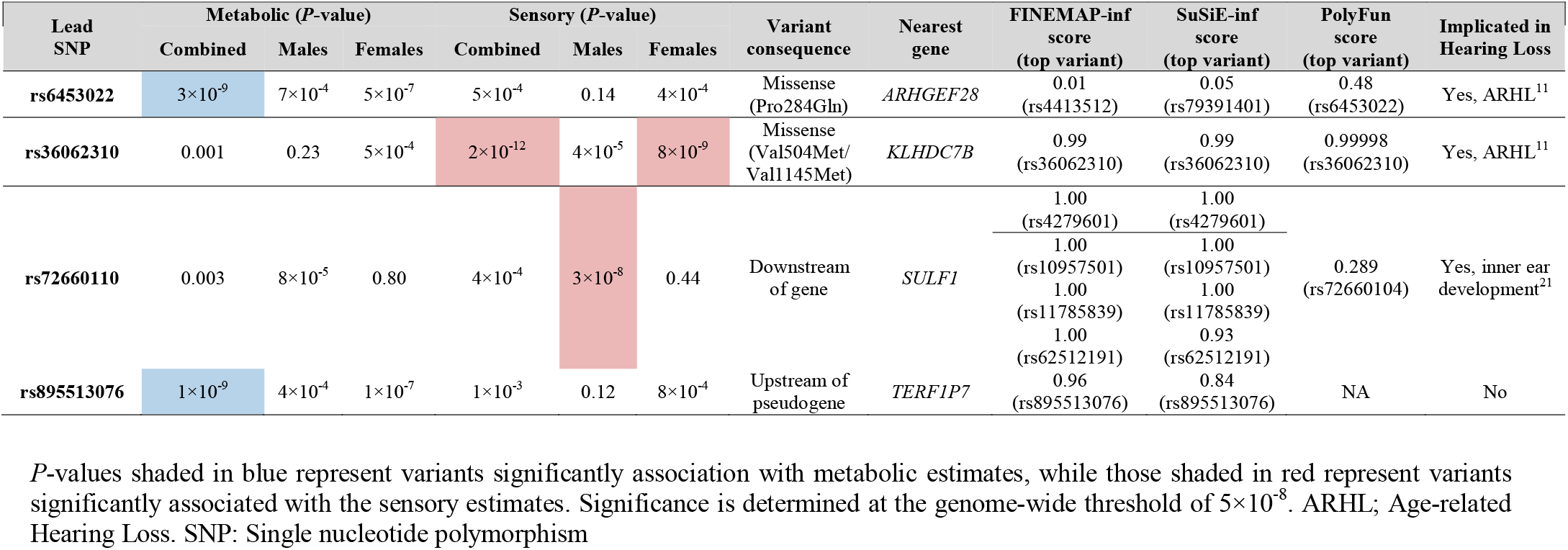
Top genomic risk loci significantly associated with metabolic and sensory hearing loss phenotypes.

### 2.5. Comparison of metabolic and sensory associations

We compared the scaled effect sizes of the genetic variants that were identified by our PolyFun analyses for both hearing loss phenotypes. Our results indicate that although the direction of effect is relatively consistent across both phenotypes, variants confer larger effects on the phenotype that they were associated with in the original GWAS analyses **(Figure 3)**. This difference in effect size is particularly pronounced for variants associated with the sensory phenotype, suggesting that genetic variants play a more significant role in susceptibility to sensory hearing loss compared to metabolic hearing loss **(Figure 3)**. These findings are in alignment with our heritability analysis results, which revealed that sensory estimates showed higher heritability estimates (*h*^*2*^*=*0.11, *P=*1.37×10^−11^) when compared to metabolic estimates (*h*^*2*^ = 0.08, *P=*9.03×10^−7^).

**Figure 3:**
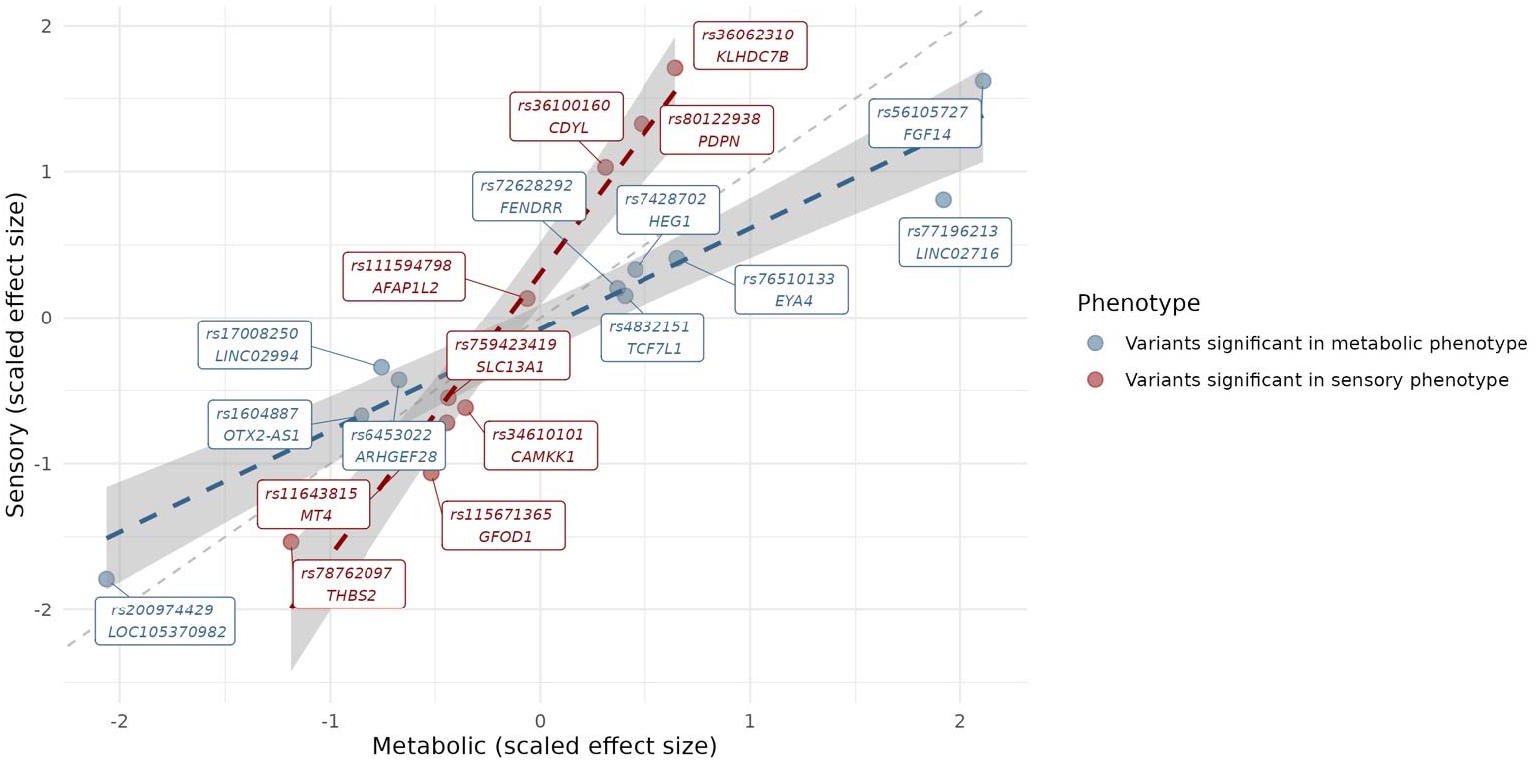
Linear regression model comparing the scaled effect sizes of variants associated with metabolic and sensory hearing loss phenotypes. The dashed grey line represents the null value of slope of 1. Shaded regions represent the 95% (CI) for combined samples. Effect sizes of variants that were significantly associated with metabolic estimates, shown in blue, have a relatively consistent direction of effect for both hearing loss phenotypes (slope: 0.69; 95% CI: 0.56-0.83; *P*=2.9 x10^−6^). Similarly, effect sizes of variants that were significantly associated with sensory estimates, shown in red, also have a relatively consistent direction of effect for both hearing loss phenotypes (slope: 1.93; 95% CI: 1.55-2.31; *P*=6.1 x10^−6^).

### 2.6. Enrichment of genetic variants in Mendelian hearing loss genes

Investigation of the GWAS results revealed that half of the genes that were significantly associated with sensory hearing loss have previously been reported to cause autosomal recessive nonsyndromic hearing loss. To investigate this further, we used a quantile-quantile (QQ) plot to visualize observed vs. expected *P* values for variants occurring in Mendelian hearing loss genes in comparison to other genes (**Figure 4)**. These analyses revealed that while variants in Mendelian hearing loss genes were enriched in both hearing loss phenotypes, this effect was more pronounced for the sensory hearing loss phenotype.

**Figure 4:** QQ plots of observed versus expected -log10(p-values) for variants associated with sensory and metabolic hearing loss phenotypes. The left plot shows results for the metabolic phenotype, and the right plot shows results for the sensory phenotype. Blue and red dots represent variants within all genes (*n*=19,438, including the X chromosome) for the metabolic and sensory phenotype, respectively, and grey dots represent variants within Mendelian hearing loss genes (*n*=193).

To investigate whether variants in the same Mendelian deafness genes were driving the observed enrichments, we extracted all variants occurring within Mendelian hearing loss genes that were associated (*P*<1×10^−3^ i.e., P<0.05 after correction for multiple testing of 193 Mendelian hearing loss genes) with either the metabolic or sensory hearing loss phenotypes. While variants in certain Mendelian hearing loss genes were associated with both phenotypes (*PCDH15, TSPEAR, ESPN, GPR98 HOMER2* and *REST*), most variants in these Mendelian hearing loss genes were only associated with either the metabolic (*n*=29 genes) or sensory (*n*=19 genes) phenotype.

Pathway analyses with EnrichR revealed that Mendelian hearing loss genes that were associated with the sensory phenotype were more likely to be implicated in pathways involved in the sensory processing of sound (*P* = 1.51×10^−10^, OR = 113.85), potassium cycling in noise-induced hearing loss (7.403×10^−10^, OR=311.86), gap junction trafficking (*P* = 8.28×10^−5^, OR = 123.68), and hair cell stereocilia protein dysfunction resulting in both congenital (*P*=4.95×10^−18^; OR=415.58) and age-related (*P*=6.03×10^−13^; OR=573.13) hearing loss. Pathways that were more significantly enriched in the metabolic phenotype included collagen synthesis (*P*=0.002, OR=45.56), and processes related to general tissue maintenance and structural integrity. A detailed list of variants and mapped genes is provided in **Supplementary Table 1**.

## 3. Discussion

By applying the novel approach developed by Vaden *et al*.^10^ to calculate metabolic and sensory hearing loss estimates from individual audiograms we were able to uncover unique genetic pathways underlying two distinct hearing loss phenotypes. The findings delineated for the first time the role of genes that have previously been associated with self-reported hearing loss in specific hearing loss phenotypes. Further, through the inclusion of specialized X-chromosome, gene-based and sex-stratified analyses, we were able to uncover novel associations between biologically plausible genes and metabolic and sensory components of ARHL.

Our GWAS analyses revealed that two missense variants, rs6453022 (p.Pro284Gln) and rs7714670 (p.Trp225Arg), in *ARHGEF28* were associated with the metabolic hearing loss phenotype. Although these variants have previously been reported to be associated with self-reported hearing difficulties **(Supplementary Table 2)**,^15^ this is the first study to define their specific role in metabolic hearing loss. While the precise mechanism through which this gene contributes to ARHL remains unclear, previous research has indicated its potential involvement in the regulation of neurofilaments, as well as axon growth and branching.^22^ As *ARHGEF28* is expressed in both inner ear hair cells and spiral ganglion neurons, its dysregulation could disrupt the transmission of electrical signals to the brain.^15^ Additionally, our gene-based approach identified two more genes, *FUS* and *IPO7*, which have not been previously associated with ARHL. Interestingly, together with *ARHGEF28* these two genes have been implicated in amyotrophic lateral sclerosis and fronto-temporal dementia.^23–25^ The previously reported role of these variants in dementia-related phenotypes adds to the growing body of evidence that shared genetic pathways may play a role in both hearing loss and cognitive decline. This is of particular interest given that hearing loss has been reported to be one of the largest modifiable risk factors for dementia.^26,27^ Our study has revealed for the first time that genes involved with metabolic hearing loss, rather than sensory hearing loss, may be driving these observed associations.

Investigation of sensory hearing loss also uncovered a significant association between a missense variant, rs36062310 (p.Val504Met), in *KLHDC7B*, which has also been strongly associated with self-reported ARHL in previous studies.^15–17,28^ Once again, this is the first study to uncover the specific importance of this gene to sensory hearing loss. While the exact mechanism by which *KLHDC7B* influences ARHL is still under investigation, ongoing research suggests that it plays a crucial role in toxin mediated apoptosis and maintaining cochlear hair cells.^20,29^ A prominent theory regarding the genetics underlying ARHL points to the involvement of genetic variants in Mendelian hearing loss genes.^30^ In line with this, our gene-based analysis identified *MYO15A* and *CLIC5* as significant contributors to sensory hearing loss. These genes are implicated in autosomal recessive non-syndromic hearing loss (DFNB3 and DFNB103, respectively).^31,32^

Broader investigation of the importance of Mendelian hearing loss genes to ARHL revealed that variants occurring within Mendelian hearing loss genes were more likely to be associated with both sensory and metabolic hearing loss when compared to other genes (**Figure 4**). With specific reference to sensory hearing loss, these Mendelian hearing loss genes were enriched in pathways related to sensory processing of sound by inner and outer hair cells. This is in alignment with reports that sensory hearing loss is typically accompanied by damage to, or death of, cochlear hair cells.^8^ While our results demonstrated that different risk loci were identified for metabolic or sensory hearing loss, it should be noted that the direction of effect for the majority of these genetic variants was consistent across both phenotypes (**Figure 3**). This could suggest that even though certain pathways may be more important for one hearing loss phenotype, the two phenotypes could also share overlapping genetic architectures. Indeed, most older adults demonstrate a combination of metabolic and sensory hearing loss,^10,33,34^ even though a negative correlation was observed between these two phenotypes (**Table 1**).

Sex-stratified GWAS analyses revealed key differences for sensory hearing loss, which is a sexually dimorphic trait. Firstly, these analyses revealed that the association between rs36062310 (p.Val1145Met; *KLHDC7B*) and sensory hearing loss was predominantly driven by the association observed in females (**Table 2**). This finding aligns with data from the International Mouse Phenotyping Consortium (IMPC) https://www.mousephenotype.org/, which reports abnormal hearing morphology in mice with *Klhdc7b* mutations in combined (male and female) and females samples, but not in males alone. Further, these analyses uncovered a new association between rs72660110, a variant mapping to *SULF1*, and sensory hearing loss in males only. Although this gene has not been previously implicated in ARHL, it has been shown to play an important role in the development of the inner ear in animal models.^21^ *SULF1* is also one of three genes that were reported to show major differences in gene expression in aged female mice compared to aged male mice.^35^ Although the exact consequences of the variants in this locus remain to be elucidated, these findings have for the first time revealed the importance of genetics in the observed sex differences for sensory hearing loss.

Previous studies have reported that the role of the X-chromosome in ARHL is limited.^36^ However, by including the X-chromosome in our analysis of hearing sub-types, we have identified a locus on the X chromosome that was associated with the metabolic phenotype. Even though annotation of this region was complicated by limited annotation information for variants occurring on the X chromosome, gene-based analyses uncovered a trend towards significance between *BCORL1* and metabolic hearing loss. This gene has previously been implicated in intellectual disability with hearing loss, and has been shown to contribute to age-related epigenetic alterations^37–39^. Therefore, this novel association warrants further investigation.

While our study provided valuable insights into the genetic pathways underlying different ARHL phenotypes, we also acknowledge several limitations. First, examination of audiograms revealed that hearing measurements taken at 0.5 kHz appear slightly elevated due to background noise during audiometric testing, as previously reported.^40^ While this limitation should be acknowledged, the overall configurations of these audiograms remain consistent with the phenotyping model that was applied to this dataset. Second, the CLSA database relies on self-reported phenotypes for many variables and contains limited information for certain demographic/clinical variables such as occupational and recreational noise exposure—key factors that influence the progression of ARHL, especially with regards to the sensory phenotype. Lastly, our cohort is predominantly composed of individuals of European ancestry (> 90%), with other populations under-represented, which may impact the generalizability of our findings to more diverse groups.

## 4. Conclusion

Our study has confirmed the heterogeneous nature of ARHL and has for the first time used GWAS to uncover specific genetic pathways that are of relevance to distinct hearing loss phenotypes. The identification of specific genes and pathways underlying metabolic and sensory hearing loss holds significant promise for the identification of drug targets for each hearing loss phenotype. For instance, genes such as *ARHGEF28* and *KLHDC7B*, which we identified as important contributors to metabolic and sensory hearing loss, respectively, exhibit high genetic priority scores (2.8 and 2.1, respectively). This is important as genes with GPS>2.1 shown to have a more than 10-fold higher chance of success in Phase IV drug development trials.^41^ These genes are also implicated in critical biological processes related to hearing, suggesting that they may be viable targets for interventions designed to prevent or mitigate ARHL. Taken together, this study has improved our understanding of the genetics underlying sensory and metabolic hearing loss and opened new avenues for future research aimed at improving early diagnosis and precise treatment of hearing loss in older adults.

## 5. Methods

### 5.1. Patient cohorts and phenotyping of hearing loss

We obtained genotype and baseline audiogram data, demographic and clinical data from 30,097 individuals included in the CLSA.^42^ Hearing thresholds were measured without the use of hearing aids at 0.5, 1, 2, 3, 4, 6 and 8 kHz by pure-tone signals ranging from 0 to 100 dB HL, with 5 dB increments, using a digital screening audiometer.^43^ Individuals with more than one missing audiogram measurement or unreliable audiometric tests were excluded from downstream analyses. Additionally, individuals were excluded if they exhibited conditions indicative of conductive hearing loss, such as the presence of ear wax, collapsed ear canals, or ear infections (**Supplementary Figure 1**). Audiometric phenotyping was determined by adopting a mathematical modeling approach developed by Vaden *et al*.^10^ This approach involved fitting individual audiogram data to previously defined hearing loss templates, which were based on average audiograms considered to be exemplars of metabolic and sensory hearing loss.^33^ In this way, we were able to obtain estimates for the extent of metabolic and sensory hearing loss for each ear in each participant. Audiograms that are well-approximated by a combination of metabolic and sensory templates had line fit error values < 15 dB (i.e., low predicted error), and were selected for inclusion in downstream analyses. Further, for each individual, we identified ears with better and worse hearing based on the calculated sensory and metabolic estimates. This study was approved by the local ethics committee.

### 5.2. Genetic data and heritability analyses

Genotype data were generated using the Affymetrix Axiom array and all samples underwent quality control (QC) and imputation using the TOPMed reference panel as previously described.^43^ Samples with call rates≥95% were included in the analyses and related individuals (pairwise kinship coefficient > 0.125), as well as those with extreme heterozygosity, genotype missingness, and discordant genetic and self-reported sex, were excluded as previously described.^43^ For marker-based QC, variants were removed based on the following criteria: variant call rates≤95%, imputation quality (R^2^)<0.8, minor allele frequency (MAF)<0.05, and Hardy-Weinberg Equilibrium (HWE) *P*<1×10^−6^ (**Supplementary Figure 2**). Samples and markers that passed genotype and phenotype QC were included to estimate SNP-based heritability for better and worse hearing ears using GCTA-GREML.^44^ For both metabolic and sensory estimates, the ear of each participant with the better hearing showed higher heritability (*h*^*2*^=0.08, *P=*9.03×10^−7^ and *h*^*2*^*=*0.11, *P=*1.37×10^−11^, respectively), compared to the ear with worse hearing (*h*^*2*^=0.058, *P=*4.49×10^−4^ and *h*^*2*^=0.105, *P=*1.19×10^−10^, respectively). Therefore, the better ear for each participant was used in all downstream genetic analyses. Better-ear metabolic and sensory estimates were normalized using the rank function in the base R Statistics software (v4.3.2)^45^.

### 5.3. Clinical and genome-wide association analyses

Linear regression was used to identify demographic and clinical variables that were associated with sensory and metabolic estimates. Forward regression was subsequently performed on significant variables (*P*<0.05, after correction for multiple testing). Variables uncovered from these analyses, along with the first ten genetic principal components provided by the CLSA, were included as covariates in the downstream genomic analyses.^43^ For autosomal chromosomes, we performed two separate GWAS using linear regression to identify genetic variants that were associated with sensory or metabolic estimates using PLINK (v1.9 and v2.0).^46,47^ To examine polymorphisms in the Human Leukocyte Antigen (*HLA*) region, we converted CLSA imputed *HLA* alleles to PLINK format using the HLA Analysis Toolkit (HATK; v2.0) and included these alleles in linear regression analyses.^48^ As hearing loss has been shown to be sexually dimorphic,^49^ we conducted sex-stratified association testing for each hearing loss estimate. Finally, we examined the association between genetic variants on the X chromosome and metabolic and sensory estimates using chromosome X-Wide Analysis toolSet (XWAS).^50^ This tool applies sex-stratified QC steps, i.e., missingness, MAF and HWE (**Supplementary Figure 2**) to account for differences in variant calling in males and females. We conducted the XWAS analyses using two models. The first model assumed complete X-inactivation, where hemizygous males were coded as having 0 or 2 alleles, corresponding to homozygous females. The second model assumed escape from X-inactivation, where males were coded as having 0 and 1 alleles corresponding to heterozygous females. *P*<5×10^−8^ was considered genome-wide significant in all SNP-based analyses. Miami plots were generated using ‘miamiplot v1.1.0’ R package.

### 5.4. Fine-mapping analyses

To uncover likely causal variants within each of the identified genomic risk loci, we defined each region by taking the lead variant flanked by 50,000 base pairs. We then implemented statistical fine-mapping analyses using Sum of Single Effects-inf (SuSiE-inf) and FINEMAP-inf methods to prioritize the variants which are most likely to be causal within each region.^51,52^ Variants with PIP>0.5 from both methods were considered likely causal. These fine-mapping analyses were further enhanced by including functional annotation using POLYgenic FUNctionally informed fine-mapping (PolyFun). Variants meeting the suggestive level of significance (*P*<1×10^−5^) were included in these analyses.^53^ We then compared the effect of these variants between metabolic and sensory associations by fitting a linear regression model between the scaled effect sizes of the variants in either of the two phenotypes at a 95% confidence interval (CI). Variants uncovered from these analyses were annotated using Ensembl’s Variant Effect Predictor (VEP)^54^ and the Combined Annotation Dependent Depletion (CADD) scoring system,^55^ using default parameters.

### 5.5. Gene and enrichment analyses

To examine the combined effects of variants within a genomic region, we performed gene-based association testing for the autosomal chromosomes using Multi-marker Analysis of GenoMic Annotation (MAGMA), implemented through FUMA.^56^ For the X-chromosome, gene-based association analyses were performed using the XWAS tool, using a modified versatile gene-based association study (VEGAS) framework.^50,57^ Genes that reached genome-wide significance (*P*<2.57×10^− 6^ i.e., adjusted for 19,438 genes) were considered significant.

Several Mendelian hearing loss genes were found to be associated with the hearing loss phenotypes included in our analyses. We therefore compared the enrichment of GWAS variants located within 50 kb of Mendelian hearing loss genes (recorded in the Hereditary Hearing Loss Database https://hereditaryhearingloss.org/)^16,17^ and non-Mendelian genes. This comparison aimed to investigate whether these hearing loss variants were more likely to be associated with sensory or metabolic hearing loss. Pathway analyses were performed using ErichR^58^ to identify specific processes related to the Mendelian hearing loss genes associated with sensory or metabolic estimates.

## 6. Acknowledgements and funding

This research was made possible using the data/biospecimens collected by the Canadian Longitudinal Study on Aging (CLSA). Funding for the CLSA is provided by the Government of Canada through the Canadian Institutes of Health Research (CIHR) under grant reference: LSA 94473 and the Canada Foundation for Innovation, as well as the following provinces, Newfoundland, Nova Scotia, Quebec, Ontario, Manitoba, Alberta, and British Columbia. This research has been conducted using the CLSA Baseline Comprehensive Dataset Version 6.0 and Genome-wide Genetic Data Version 3.0 under Application Number 2104035. The CLSA is led by Drs. Parminder Raina, Christina Wolfson and Susan Kirkland. The opinions expressed in this manuscript are the author’s own and do not reflect the views of the Canadian Longitudinal Study on Aging.

This work was funded through a Natural Sciences and Engineering Research Council of Canada Discovery Grant (B.D.) and (in part) by the National Institutes of Health/National Institute on Deafness and Other Communication Disorders Clinical Research Center (P50 DC 000422) awarded to the Medical University of South Carolina and by the South Carolina Clinical and Translational Research (SCTR) Institute, with an academic home at the Medical University of South Carolina, NIH/NCATS Grant number UL1 TR001450 (J.R.D. and K.V.). Portions of this investigation were conducted in a facility constructed with support from Research Facilities Improvement Program Grant Number C06 RR14516 from the NIH/NCRR (J.R.D. and K.V.). BD is supported by a CIHR Tier 2 Canada Research Chair in Pharmacogenomics and Precision Medicine. SA was supported through a Research Manitoba Studentship and the Visual and Automated Disease Analytics Graduate Training Program at the University of Manitoba.

## 7. Author contributions

S.A. performed the analyses and wrote the manuscript. K.I.V. and J.R.D developed the phenotyping approach, provided clinical expertise and edited the manuscript, D.L. provided clinical expertise and edited the manuscript, B.I.D. conceived of and supervised the project and edited the manuscript.

## 9. Data availability

Data are available from the Canadian Longitudinal Study on Aging (www.clsa-elcv.ca) for researchers who meet the criteria for access to de-identified CLSA data. Scripts that were used in this study are available on https://github.com/Drogemoller-Lab/.

## Reference

1. Deafness and hearing loss. https://www.who.int/news-room/fact-sheets/detail/deafness-and-hearing-loss.

2. Wattamwar, K. et al. Increases in the Rate of Age-Related Hearing Loss in the Older Old. JAMA Otolaryngol Head Neck Surg 143, 41–45 (2017).

3. Prince, M. J. et al. The burden of disease in older people and implications for health policy and practice. The Lancet 385, 549–562 (2015).

4. Kotby, M. N., Tawfik, S., Aziz, A. & Taha, H. Public health impact of hearing impairment and disability. Folia Phoniatr Logop 60, 58–63 (2008).

5. Tsimpida, D., Kontopantelis, E., Ashcroft, D. & Panagioti, M. Comparison of Self-reported Measures of Hearing With an Objective Audiometric Measure in Adults in the English Longitudinal Study of Ageing. JAMA Netw Open 3, e2015009 (2020).

6. Yamasoba, T. et al. Current concepts in age-related hearing loss: Epidemiology and mechanistic pathways. Hear Res 303, 30–38 (2013).

7. Lewis, M. A. et al. Accurate phenotypic classification and exome sequencing allow identification of novel genes and variants associated with adult-onset hearing loss. PLoS Genet 19, e1011058 (2023).

8. Wu, P., O’Malley, J. T., Gruttola, V.de & Liberman, M. C. Age-Related Hearing Loss Is Dominated by Damage to Inner Ear Sensory Cells, Not the Cellular Battery That Powers Them. J. Neurosci. 40, 6357–6366 (2020).

9. Schmiedt, R. A. Effects of aging on potassium homeostasis and the endocochlear potential in the gerbil cochlea. Hear Res 102, 125–132 (1996).

10. Vaden, K. I., Eckert, M. A., Matthews, L. J., Schmiedt, R. A. & Dubno, J. R. Metabolic and Sensory Components of Age-Related Hearing Loss. JARO 23, 253–272 (2022).

11. Karlsson, K. K., Harris, J. R. & Svartengren, M. Description and primary results from an audiometric study of male twins. Ear Hear 18, 114–120 (1997).

12. Gates, G. A., Couropmitree, N. N. & Myers, R. H. Genetic associations in age-related hearing thresholds. Arch Otolaryngol Head Neck Surg 125, 654–659 (1999).

13. Wingfield, A. et al. A Twin-Study of Genetic Contributions to Hearing Acuity in Late Middle Age. J Gerontol A Biol Sci Med Sci 62, 1294–1299 (2007).

14. Duan, H. et al. Heritability of Age-Related Hearing Loss in Middle-Aged and Elderly Chinese: A Population-Based Twin Study. Ear and Hearing 40, 253 (2019).

15. Wells, H. R. R. et al. GWAS Identifies 44 Independent Associated Genomic Loci for Self-Reported Adult Hearing Difficulty in UK Biobank. Am J Hum Genet 105, 788–802 (2019).

16. Kalra, G. et al. Biological insights from multi-omic analysis of 31 genomic risk loci for adult hearing difficulty. PLOS Genetics 16, e1009025 (2020).

17. Liu, W., Johansson, Å., Rask-Andersen, H. & Rask-Andersen, M. A combined genome-wide association and molecular study of age-related hearing loss in H. sapiens. BMC Medicine 19, 302 (2021).

18. Nolan, L. S. Age-related hearing loss: Why we need to think about sex as a biological variable. Journal of Neuroscience Research 98, 1705–1720 (2020).

19. Mick, P. T. et al. The Prevalence of Hearing, Vision, and Dual Sensory Loss in Older Canadians: An Analysis of Data from the Canadian Longitudinal Study on Aging. Can J Aging 40, 1–22 (2021).

20. Yahiro, K. et al. A novel endoplasmic stress mediator, Kelch domain containing 7B (KLHDC7B), increased Harakiri (HRK) in the SubAB-induced apoptosis signaling pathway. Cell Death Discov. 7, 1–11 (2021).

21. Freeman, S. D., Keino-Masu, K., Masu, M. & Ladher, R. K. Expression of the heparan sulfate 6-O-endosulfatases, Sulf1 and Sulf2, in the avian and mammalian inner ear suggests a role for sulfation during inner ear development. Dev Dyn 244, 168–180 (2015).

22. Volkening, K., Leystra-Lantz, C. & Strong, M. J. Human low molecular weight neurofilament (NFL) mRNA interacts with a predicted p190RhoGEF homologue (RGNEF) in humans. Amyotroph Lateral Scler 11, 97–103 (2010).

23. Neumann, M. et al. A new subtype of frontotemporal lobar degeneration with FUS pathology. Brain 132, 2922–2931 (2009).

24. Katsumata, Y. et al. Genetic associations with dementia_Jrelated proteinopathy: Application of item response theory. Alzheimers Dement 20, 2906–2921 (2024).

25. Kim, W.Kim, D.-Y. & Lee, K.-H. RNA-Binding Proteins and the Complex Pathophysiology of ALS. Int J Mol Sci 22, 2598 (2021).

26. Son, S. et al. Potentially Modifiable Dementia Risk Factors in Canada: An Analysis of Canadian Longitudinal Study on Aging with a Multi-Country Comparison. J Prev Alzheimers Dis (2024) doi:10.14283/jpad.2024.105.

27. Livingston, G. et al. Dementia prevention, intervention, and care: 2024 report of the Lancet standing Commission. Lancet 404, 572–628 (2024).

28. Trpchevska, N. et al. Genome-wide association meta-analysis identifies 48 risk variants and highlights the role of the stria vascularis in hearing loss. Am J Hum Genet 109, 1077–1091 (2022).

29. Kaufman, A. Exploring the Function of KLHDC7b, a Novel Gene Associated With Hearing Loss. in (Anaheim, CA, USA, 2024).

30. Ivarsdottir, E. V. et al. The genetic architecture of age-related hearing impairment revealed by genome-wide association analysis. Commun Biol 4, 1–13 (2021).

31. Wang, A. et al. Association of unconventional myosin MYO15 mutations with human nonsyndromic deafness DFNB3. Science 280, 1447–1451 (1998).

32. Wonkam-Tingang, E. et al. Bi-Allelic Novel Variants in CLIC5 Identified in a Cameroonian Multiplex Family with Non-Syndromic Hearing Impairment. Genes (Basel) 11, 1249 (2020).

34. Schmiedt, R. A. The Physiology of Cochlear Presbycusis. in The Aging Auditory System (eds. Gordon-Salant, S., Frisina, R. D., Popper, A.N. & Fay, R.R.) 9–38 (Springer, New York, NY, 2010). doi:10.1007/978-1-4419-0993-0_2.

35. Wang, Y. et al. Sex differences in transcriptomic profiles in aged kidney cells of renin lineage. Aging (Albany NY) 10, 606–621 (2018).

36. Naderi, E. et al. The genetic contribution of the X chromosome in age-related hearing loss. Front. Genet. 14, (2023).

37. Barone, C. et al. Intragenic ILRAPL1 deletion in a male patient with intellectual disability, mild dysmorphic signs, deafness, and behavioral problems. Am J Med Genet A 161A, 1381–1385 (2013).

38. Shukla, A. et al. Variants in the transcriptional corepressor BCORL1 are associated with an X-linked disorder of intellectual disability, dysmorphic features, and behavioral abnormalities. Am J Med Genet A 179, 870–874 (2019).

39. Wonkam, A. et al. Exome sequencing of families from Ghana reveals known and candidate hearing impairment genes. Commun Biol 5, 369 (2022).

40. Mick, P. T. et al. Associations Between Cardiovascular Risk Factors and Audiometric Hearing: Findings From the Canadian Longitudinal Study on Aging. Ear and Hearing 44, 1332 (2023).

41. Duffy, Á. et al. Development of a human genetics-guided priority score for 19,365 genes and 399 drug indications. Nat Genet 56, 51–59 (2024).

42. Raina, P. et al. Cohort Profile: The Canadian Longitudinal Study on Aging (CLSA). International Journal of Epidemiology 48, 1752–1753j (2019).

43. Forgetta, V. et al. Cohort profile: genomic data for 26 622 individuals from the Canadian Longitudinal Study on Aging (CLSA). BMJ Open 12, e059021 (2022).

44. Yang, J., Lee, S. H., Goddard, M. E. & Visscher, P. M. GCTA: a tool for genome-wide complex trait analysis. Am J Hum Genet 88, 76–82 (2011).

45. R Core Team (2023). R: A Language and Environment for Statistical Computing_. R Foundation for Statistical Computing, Vienna, Austria. <https://www.R-project.org/>.

46. Purcell, S. et al. PLINK: A Tool Set for Whole-Genome Association and Population-Based Linkage Analyses. Am J Hum Genet 81, 559–575 (2007).

47. Purcell, S. PLINK v1.09, v2.0. http://pngu.mgh.harvard.edu/purcell/plink/.

48. Choi, W., Luo, Y., Raychaudhuri, S. & Han, B. HATK: HLA analysis toolkit. Bioinformatics 37, 416–418 (2021).

49. De Angelis, F. et al. Sex differences in the polygenic architecture of hearing problems in adults. Genome Med 15, 1–18 (2023).

50. Gao, F. et al. XWAS: A Software Toolset for Genetic Data Analysis and Association Studies of the X Chromosome. J Hered 106, 666–671 (2015).

51. Cui, R. et al. Improving fine-mapping by modeling infinitesimal effects. 2022.10.21.513123 Preprint at 10.1101/2022.10.21.513123 (2022).

52. Cui, R. et al. Improving fine-mapping by modeling infinitesimal effects. Nat Genet 56, 162–169 (2024).

53. Weissbrod, O. et al. Functionally informed fine-mapping and polygenic localization of complex trait heritability. Nat Genet 52, 1355–1363 (2020).

54. McLaren, W. et al. The Ensembl Variant Effect Predictor. Genome Biology 17, 122 (2016).

55. Kircher, M. et al. A general framework for estimating the relative pathogenicity of human genetic variants. Nat Genet 46, 310–315 (2014).

56. Leeuw, C. A. de, Mooij, J. M., Heskes, T. & Posthuma, D. MAGMA: Generalized Gene-Set Analysis of GWAS Data. PLOS Computational Biology 11, e1004219 (2015).

57. Liu, J. Z. et al. A versatile gene-based test for genome-wide association studies. Am J Hum Genet 87, 139–145 (2010).

58. Chen, E. Y. et al. Enrichr: interactive and collaborative HTML5 gene list enrichment analysis tool. BMC Bioinformatics 14, 128 (2013).

