## Supplemental Table 1 for "Large-scale audiometric phenotyping identifies distinct genes and pathways involved in hearing loss subtypes"

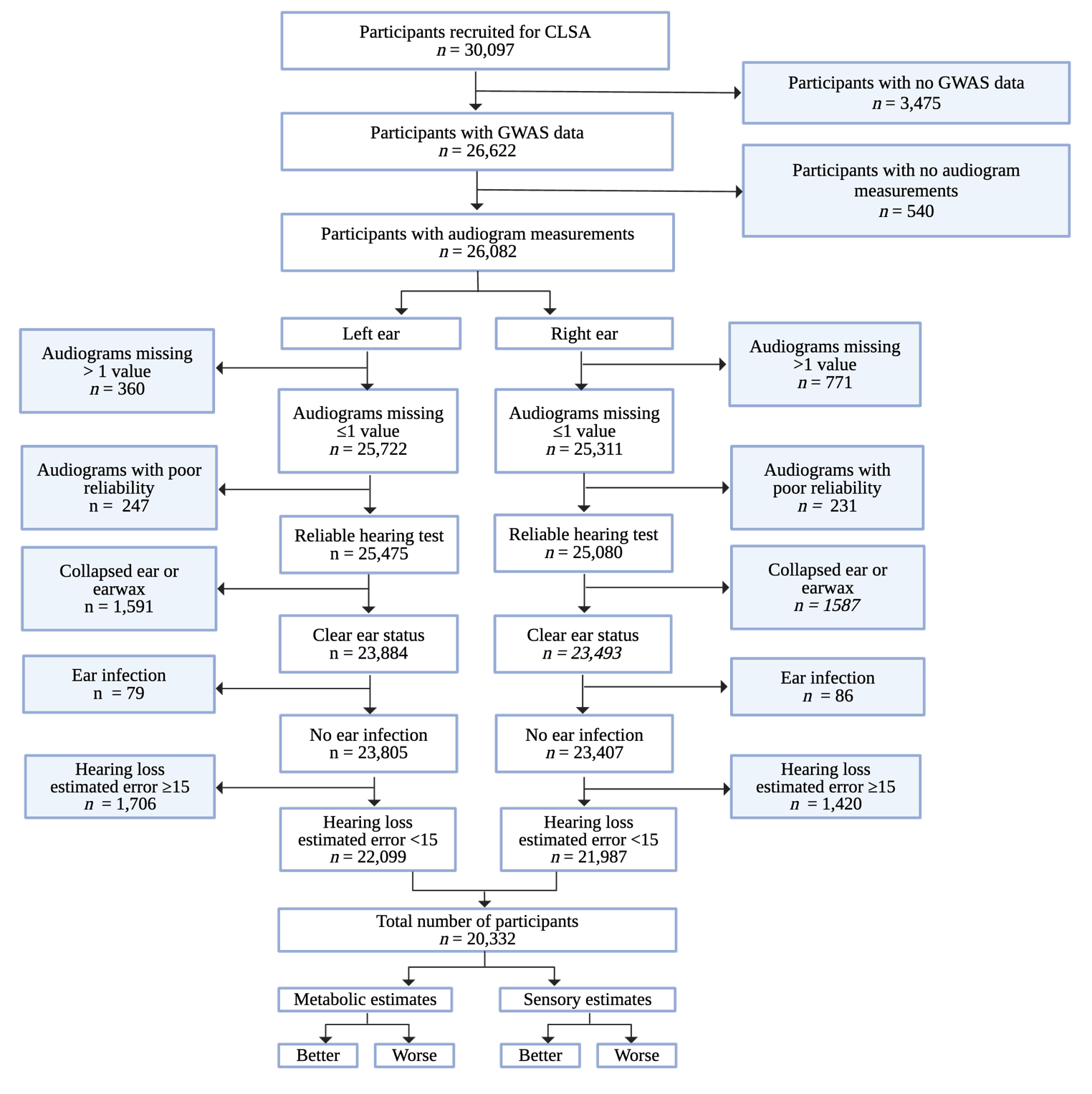

**Supplementary Figure 1: Flow diagram of audiogram data processing and selection of samples.** GWAS: Genome-Wide Association Study; CLSA: Canadian Longitudinal Study on Aging

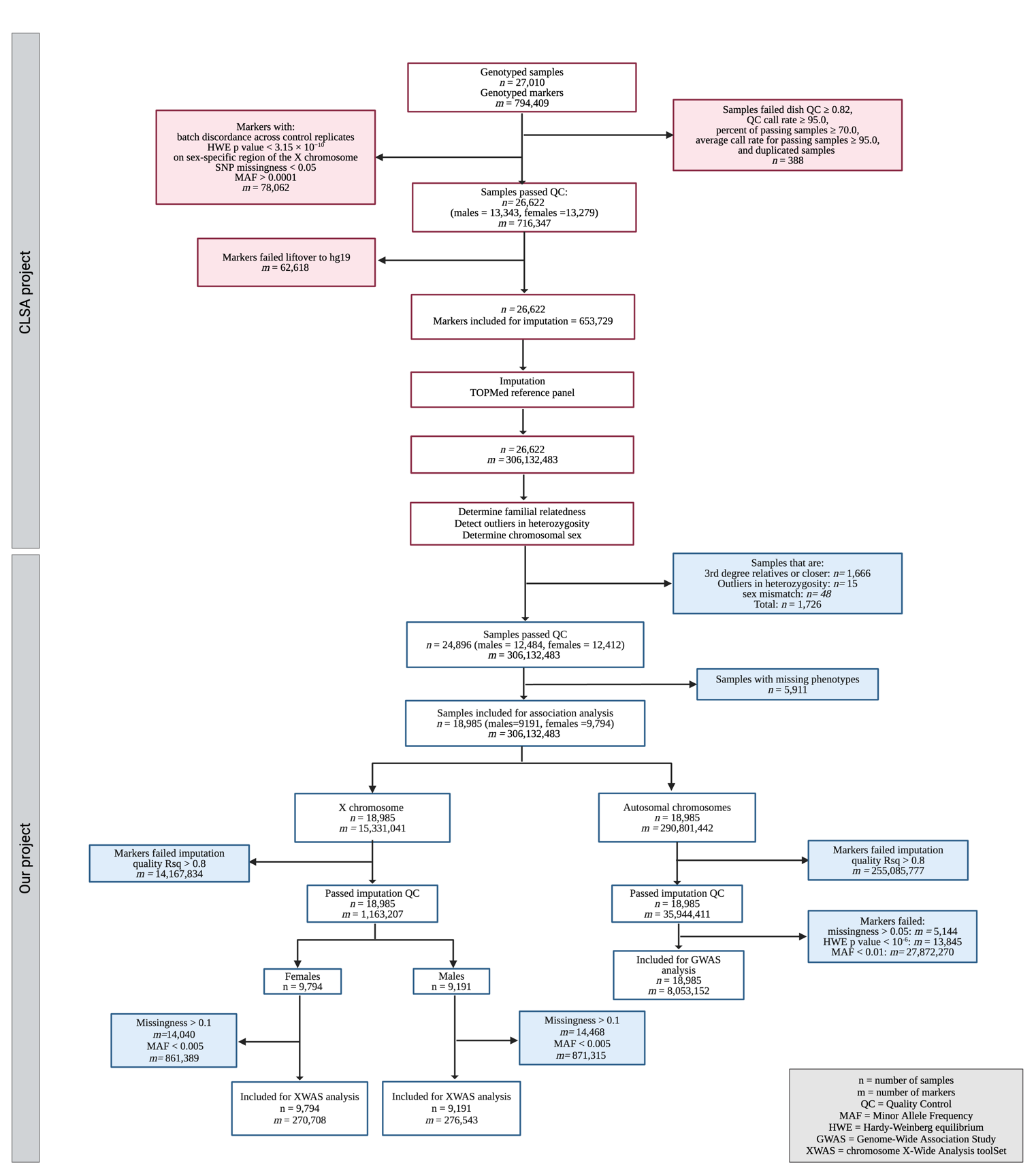

**Supplementary Figure 2: Flow diagram illustrating the quality control steps for both samples and markers in the CLSA study.** The section highlighted in red indicates steps performed in CLSA's original genomic quality control analyses, while the blue section represents steps conducted in our analysis. CLSA: Canadian Longitudinal Study on Aging.

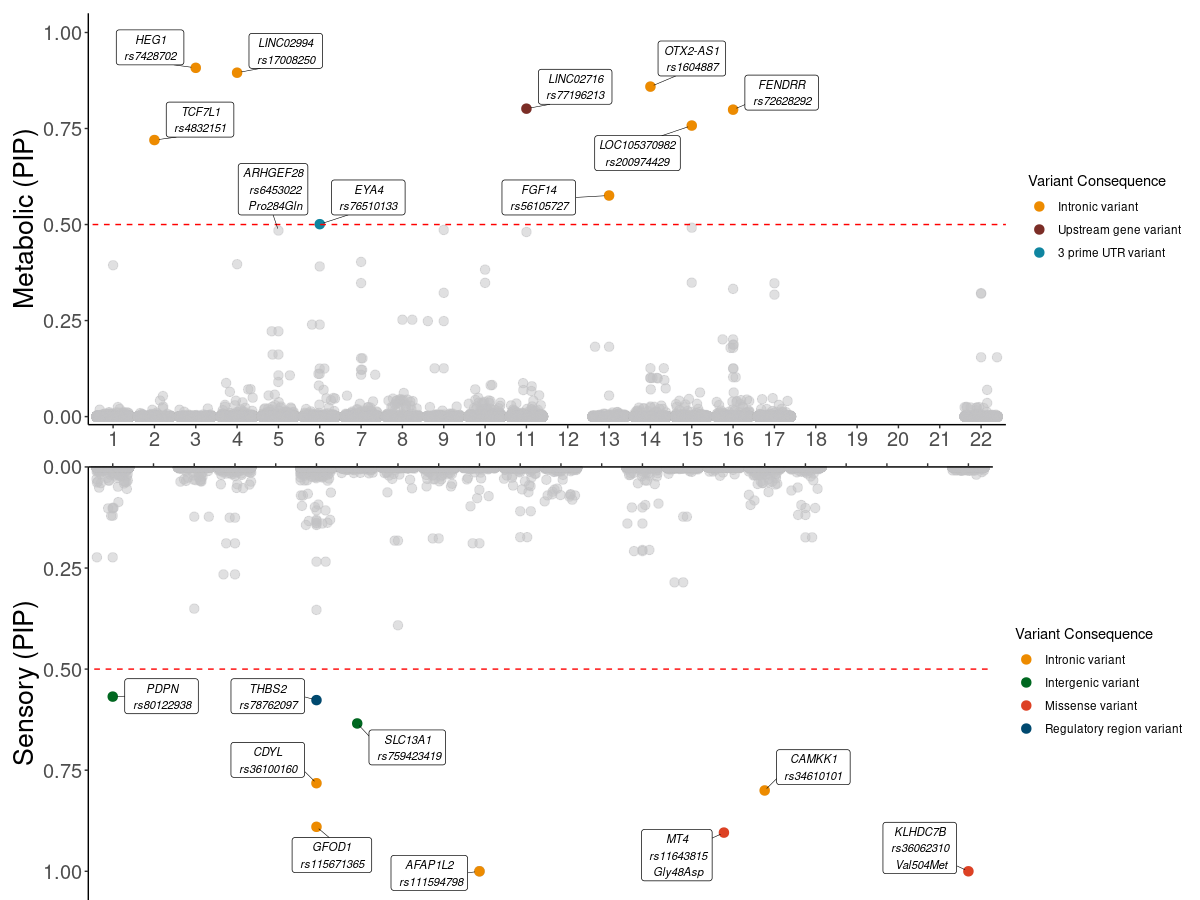

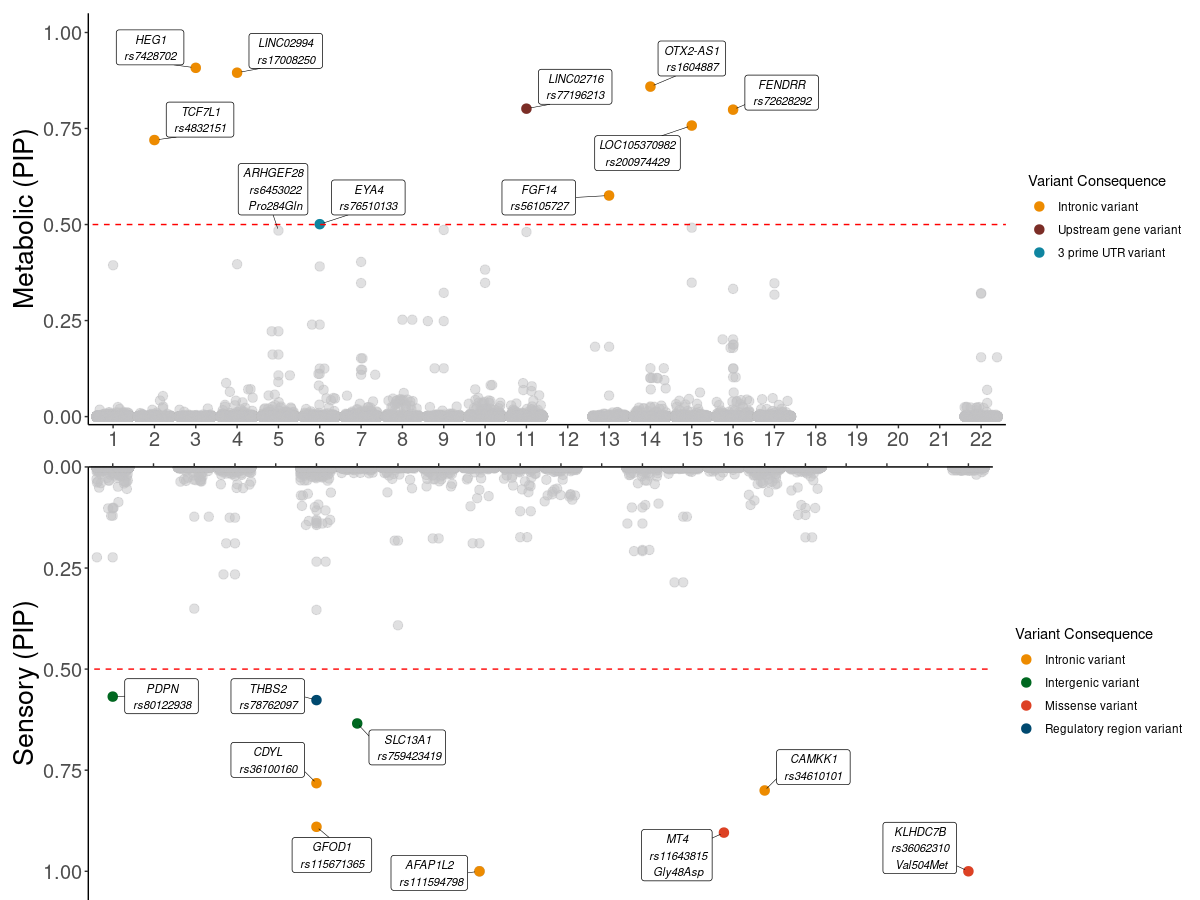

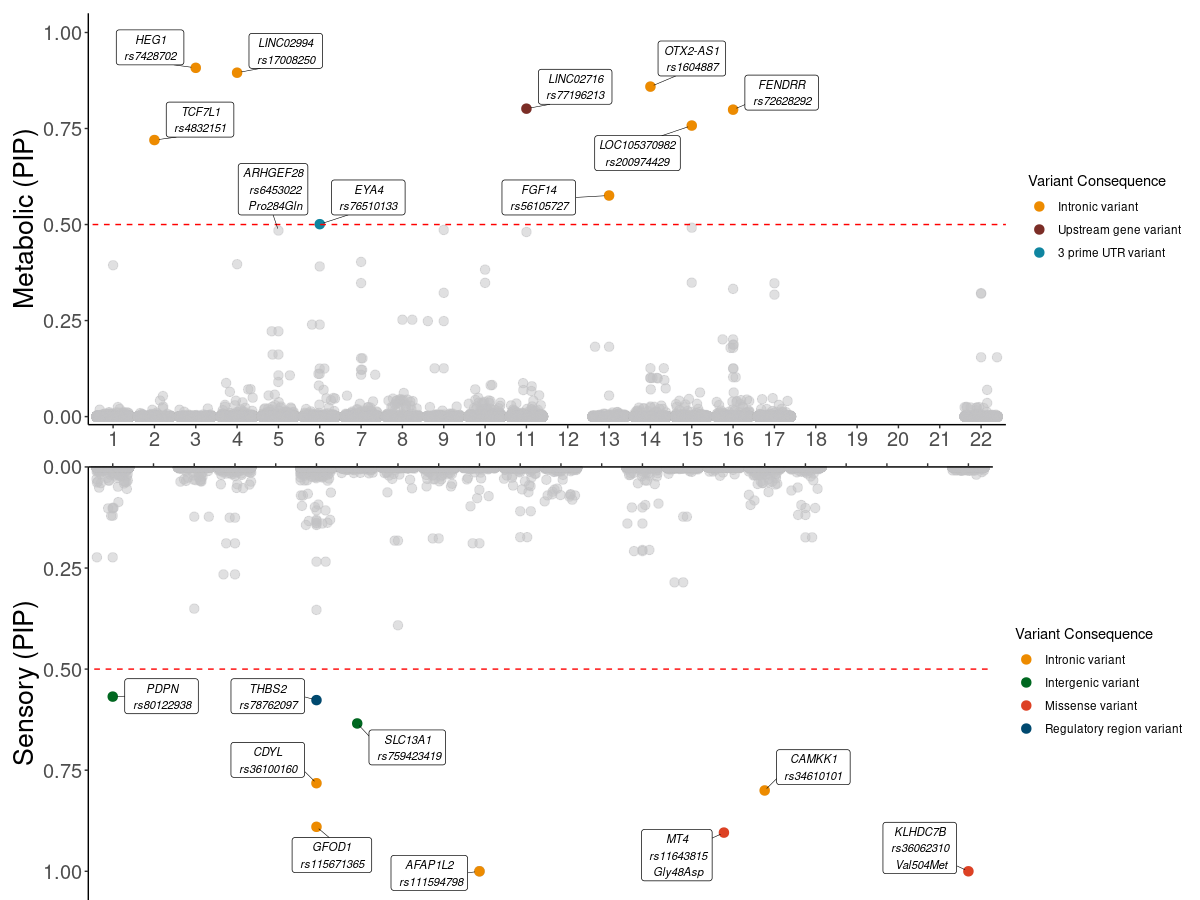

**Supplementary Figure** **3: Fine-mapping results from the PolyFun of all regions containing variants with suggestive level of significant (*P* < 1×10^-5^).** FINEMAP PIP scores are shown for metabolic and sensory phenotypes in the upper and lower panel, respectively. Significant variants are colored according to their functional consequences. The dashed red line represents PIP score threshold of 0.5. PIP: Posterior inclusion probability

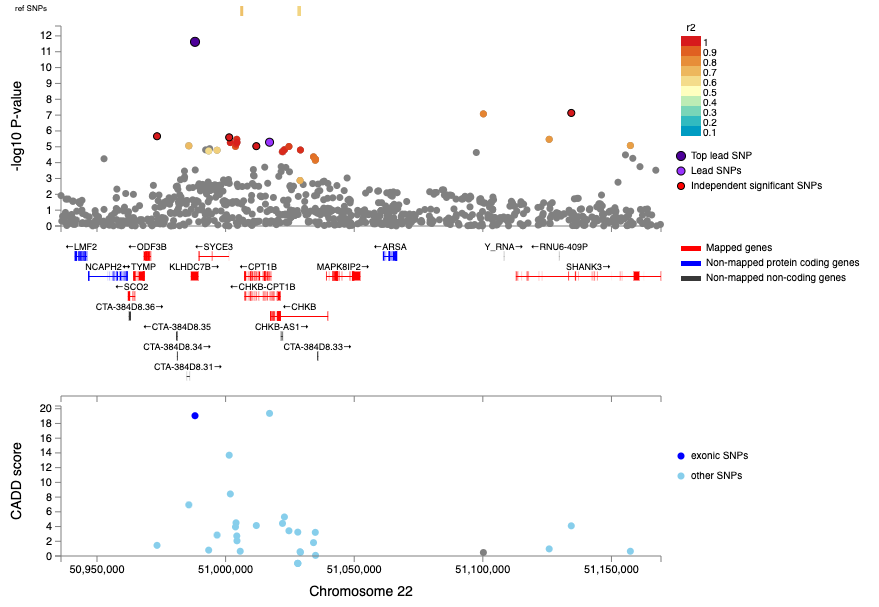

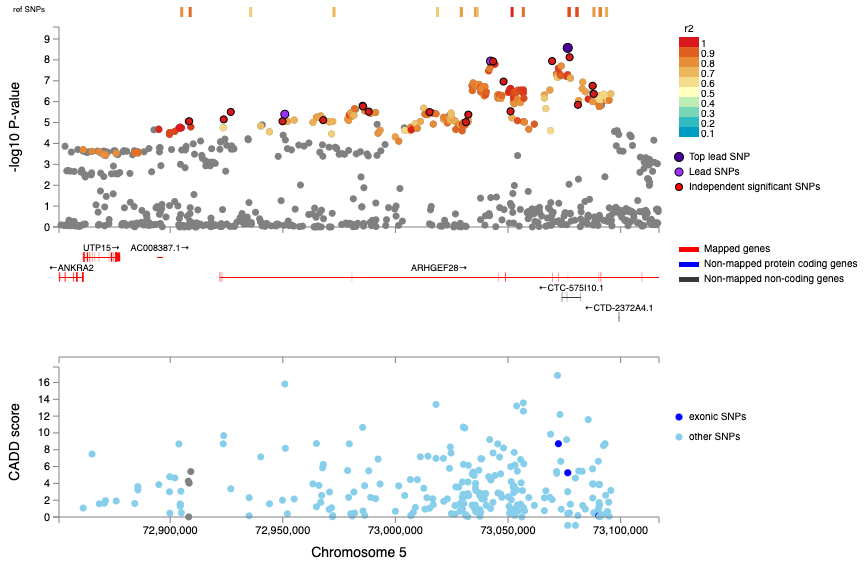

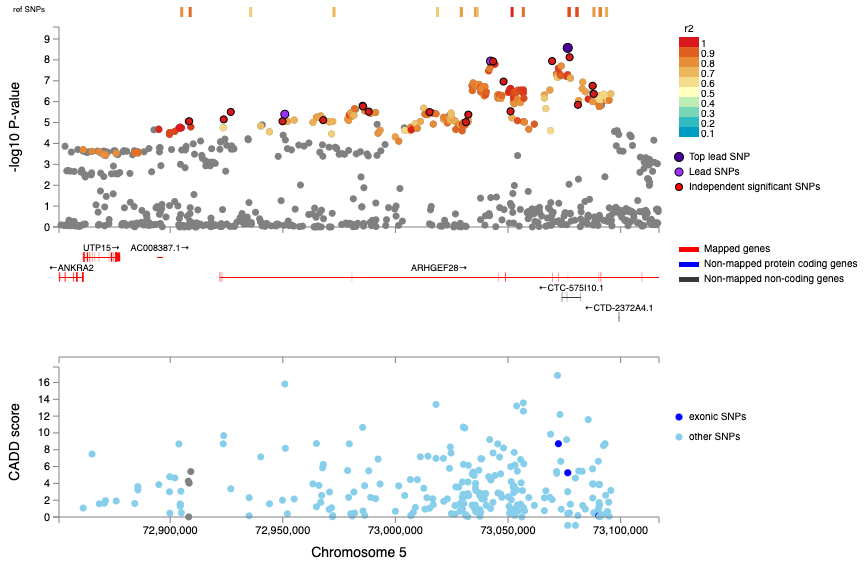

rs6453022

rs36062310

A

B

**Supplementary Figure 4**: **LocusZoom plots for risk loci significantly associated with** **(A)** the metabolic phenotype and **(B)** sensory phenotypes along with their CADD scores (lower panel). CADD; Combined Annotation Dependent Depletion.

B
